# Spatial remodeling of the urothelial carcinoma tumor microenvironment shapes response to neoadjuvant atezolizumab

**DOI:** 10.64898/2026.04.15.26350980

**Authors:** Robbin Nameki, Jennifer Kinong, Chao-Hui Huang, Michelle Saul, Aakash Sur, Arne Schmidt, Nina Kozar-Gillan, Sophie Laturnus, Mehmet Tekman, Alexander Trageser, Wenjing Yang, Daniel Chawla, Alberto Megina Gonzalo, Srishti Munjal Mehta, Rosemarie Krupar, Cornelius Böhm, Marija Pezer, Gloria H.Y. Lin, Diane Fernandez, William E. Pierceall, Jadwiga R. Bienkowska, Gregory Lee Szeto, Craig B Davis, Thomas Powles, Keith Ching

## Abstract

The ABACUS study was a single arm, phase II trial evaluating neoadjuvant atezolizumab in operable urothelial carcinoma. Initial bulk transcriptomic and immunohistochemistry analyses suggested links between immune activation, tissue remodeling, and resistance pathways such as transforming growth factor β that were associated with clinical outcome. To further characterize spatial and phenotypic changes at high resolution, artificial intelligence–assisted digital image analysis of hematoxylin and eosin sections and spatial transcriptomics were performed on paired tissue samples. In baseline samples, cells residing in lymphoid aggregates and tertiary lymphoid structures were more abundant in stable disease than in relapse and exhibited gene expression programs associated with improved survival in urothelial carcinoma. Most spatial features reflected shared pharmacodynamic changes between stable disease and relapse; however, carcinoma-endothelial adjacency was reduced significantly following treatment and differed between groups, accompanied by distinct transcriptional programs. Together, these findings indicate that atezolizumab induces localized immune and stromal remodeling within the tumor microenvironment, while non-response despite immune expansion is associated with persistent spatial immune exclusion and carcinoma-endothelial adjacency. Spatial and phenotypic biomarkers identified here may inform rational combination strategies for immune checkpoint inhibitor–refractory urothelial carcinoma.

## 1. INTRODUCTION

Immune checkpoint inhibitors (ICIs) have transformed the treatment landscape for urothelial carcinoma (UC), demonstrating a durable clinical benefit in advanced and neoadjuvant settings (1–5). Despite these advances, substantial heterogeneity in clinical response persists, and established biomarkers such as PD-L1 expression, tumor mutational burden, and CD8+ T cell abundance have shown modest ability to identify patients most likely to benefit from treatment (6–8).

The ABACUS trial evaluated neoadjuvant atezolizumab in operable UC and established the clinical activity of PD-L1 blockade in this setting (9). Translational biomarker analyses from ABACUS based on bulk transcriptomics and immunohistochemistry identified associations between the treatment outcome with pre-existing CD8+ T cell immunity, stromal components, and post-treatment CD8+ T cell phenotype. Transforming Growth Factor-beta (TGFβ) signatures and fibroblast activation protein (FAP) were enriched in immune excluded samples and non-responders, similarly to the metastatic setting where infiltrating T cells were previously observed to be enriched in fibroblast-rich peritumoral stroma (10). These findings suggest that response to PD-L1 blockade reflects complex, spatially coordinated remodeling of immune and stromal compartments.

Emerging evidence indicates that spatial context influences the outcome of immune checkpoint blockade. The tumor microenvironment is spatially organized into structured niches that drive distinct biology, including immune effector cell proximity to tumor cells to drive killing, immune exclusion driven by stromal-T cell interactions, and T and B cell priming in mature TLS (11, 12). Multiple studies report that these spatially resolved biomarkers are characterized by distinct cellular composition, phenotype and location which can be difficult to infer from bulk measurements and can be associated with clinical outcomes independently from traditional biomarkers like PD-L1 expression and CD8+ T cell abundance (12).

Artificial intelligence–based digital image analysis (DIA) of histopathology and spatial transcriptomics now enable systematic characterization of cellular composition and spatial relationships at high resolution (13–15). These advances provide a powerful framework for interrogating how PD-L1 blockade reshapes the tumor microenvironment and for identifying strategies to enhance therapeutic response. Here, we leverage AI based DIA with spatial transcriptomics (10x Genomics, Visium) on paired pretreatment biopsies and post-treatment resections from the ABACUS cohort to characterize treatment-associated spatial changes following neoadjuvant atezolizumab.

## 2. RESULTS

### 2.1. Cohort overview and integrated spatial profiling workflow

The ABACUS study was a single-arm phase II study to evaluate the efficacy of atezolizumab in UC at a neoadjuvant setting (9). Of the 95 patients recruited across 21 sites, 64 patients met primary endpoints with paired baseline biopsy and post-treatment cystectomy (Figure 1A). Here we examined the tumor microenvironment in 26 ABACUS patients (15 with stable disease and 11 with relapse) whose paired samples met inclusion criteria for DIA and spatial transcriptomics (DV200 > 20% and board-certified pathological assessment) (Figure 1B). Given the modest cohort size, these pharmacodynamic analyses are exploratory and intended to generate mechanistic hypotheses.

**Figure 1.**
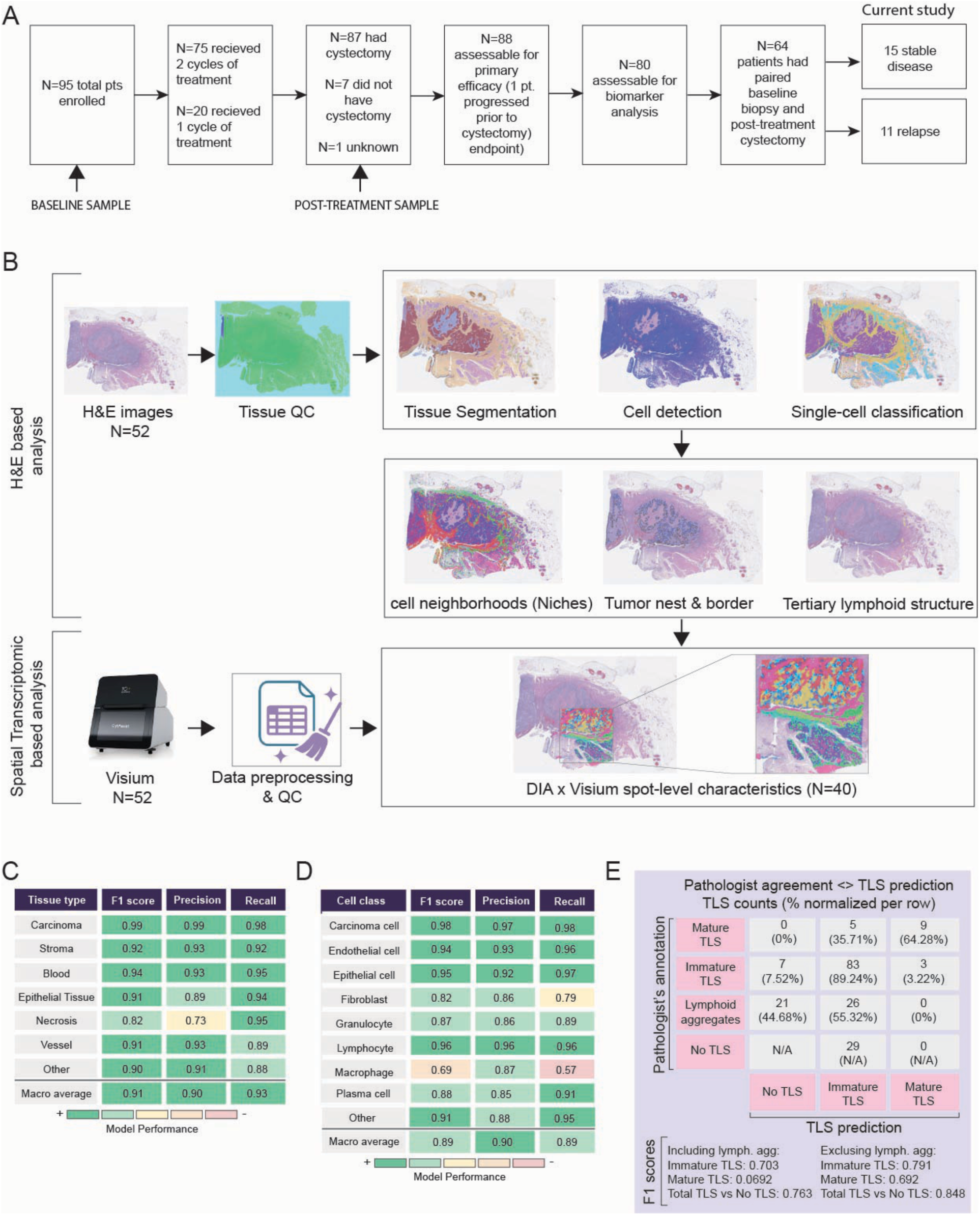
Characterization of UC tumor microenvironment by digital image analysis and spatial transcriptomics. **(A)** Study design and sample collection workflow for the ABACUS UC cohort. Patients received 1–2 cycles of atezolizumab between diagnostic biopsy and cystectomy. A total of 64 baseline and on-treatment samples were collected, with a subset of 26 paired samples passing inclusion criteria for analysis. **(B)** Integrated spatial profiling workflow. HCE whole-slide images were generated and annotated by Atlas HCE-TME classifier with a Digital Image Analysis foundation model, followed by Visium spatial transcriptomics to enable joint spatial and molecular analysis. **(C–E)** Precision, recall, and F1 performance of the Atlas HCE-TME classifier with a Digital Image Analysis foundation model for cell types, tissue segmentation, and for TLS detection using a separate TLS-specific model. Validated using annotations provided by a board-certified pathologist.

The Atlas HCE-TME DIA workflow built on Aignostics’ foundation models (16, 17) was applied in a structured sequence, beginning with tissue quality control and progressing through tissue segmentation, cell detection, and cell classification. Identification of lymphoid aggregate and tertiary lymphoid structures (LAs/TLSs) were performed separately using a dedicated TLS detection model (Figure 1B). For cell classification, T cells and non-plasma B cells were grouped as a single lymphocyte category as HCE morphology does not reliably distinguish T cells from non-plasma B cells. Plasma cells were classified separately based on their distinctive morphology. Beyond these model derived outputs, higher order spatial features were generated through distinct analytical strategies. Cellular neighborhoods were defined using unsupervised k-means clustering based on the relative abundance of classified cell types within a 50 μm radius around each cell, resulting in 25 initial niches. The 25 niches were further consolidated by a board-certified pathologist into 16 niches (Supplementary Table 1). Tumor nests and their immune borders were derived from supervised criteria applied to the carcinoma segmentation mask. These image-based features were subsequently aligned to serial section spatial transcriptomic datasets, enabling direct linkage between whole slide morphology and spot level transcriptional programs (Figure 1B). Of the 52 spatial transcriptomic samples, 40 were retained for downstream analysis following sample and spot level quality control based on gene detection and probe mapping metrics.

Tissue segmentation, cell classification, and TLS classifier performance were evaluated through a combination of pathologist review and quantitative benchmarking on a dedicated holdout set comprising approximately 20% of all cohort slides. The QC model accurately distinguished valid from invalid tissue (F1 = 0.95), and the tissue segmentation, cell classification, and TLS models achieved class averaged F1 scores of 0.91, 0.89, and 0.76, respectively. Performance for key biological classes remained high, including an F1 of 0.99 for carcinoma segmentation and F1 scores of 0.98 and 0.96 for carcinoma cells and lymphocytes, respectively (Figure 1C-E).

### 2.2. Spatial features of lymphoid aggregates and TLS in pre- and post-treatment specimens

TLS are ectopic lymphoid aggregates that support local antigen presentation and B and T cell coordination, and their baseline presence has been associated with improved survival and response to anti-PD-1/L1 therapy across multiple tumor types (12, 18, 19). Functional studies demonstrate that TLS induction enhances response to PD1 blockade, further supporting their mechanistic relevance in immunotherapy (20–22). Given this, we first examined TLS architecture and spatial distribution in ABACUS as a foundation for pharmacodynamic analysis.

At baseline, whole-slide TLS density classified by DIA did not differ significantly between stable disease (SD) and relapse groups, likely reflecting substantial inter-sample variability and limited sample size (Supplementary Fig. 1A, P = 0.59). We therefore next assessed whether spatially resolved analyses could reveal localized TLS-associated differences that are obscured by slide-level aggregation. Using the H-plot approach to assess spatial patterns relative to the tumor border, both pre- and post-treatment SD groups demonstrated an enrichment in proportion of TLS originating cells within the intermediate spatial zone spanning approximately 200–800 µm from the tumor border, compared to relapse tumors (Kruskal-Wallis test across distance layers within 200-800 µm, p = 2.7×10^-31^) . The pre-treatment relapse group exhibited the lowest proportion of TLS origination cells within this immediate peritumoral region. Beyond 800 μm, measurements became increasingly variable, suggesting predominantly regional spatial effects (Figure 2A).

**Figure 2.**
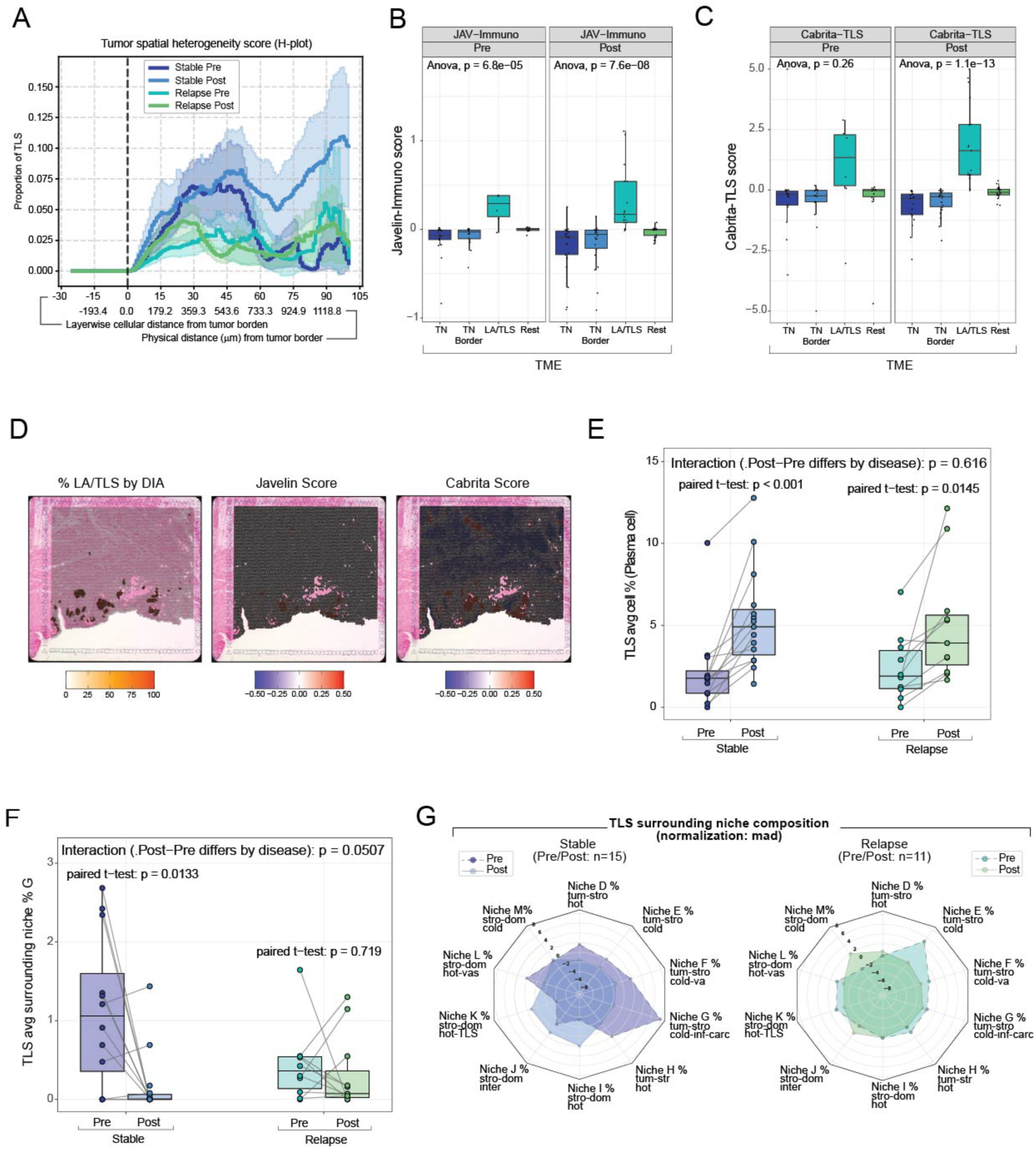
Spatial features of lymphoid aggregates and tertiary lymphoid structures in baseline and post-treatment specimens. **(A)** H-plot of TLS originating cell type proportions with 80% CI (stable disease pre n = 12, stable disease post n = 15, relapse pre n = 10, relapse post n = 11). **(B)** Visium spatial transcriptomics data of TN (Pre n = 18, Post n = 22), TN Border (Pre n = 16, Post n = 22), LA/TLS (Pre n = 6, Post n = 17), Rest (Pre n = 14, Post n = 22) represented as box and whisker plot (box spans interquartile range, center line indicates median, whiskers extend to 1.5× IQR) for JAV-Immuno and **(C)** Cabrita Seurat module scores. Statistical significance was assessed using ANOVA, with exact P values reported. **(D)** Representative feature plots of Visium spatial transcriptomics for % LA/TLS per spot by DIA and Seurat module scores for JAV-Immuno and Cabrita. Color scale represents score intensity. **(E)** DIA data for the average percent plasma cell within TLSs (stable pair n = 12, relapse pair n = 10) represented as spaghetti plots (box spans interquartile range, center line indicates median, whiskers extend to 1.5× IQR) with lines connecting paired samples. Paired differences were assessed using a paired t-test, and group-by-outcome effects were assessed using an interaction test. Exact P values are reported unless P < 0.001. **(F)** DIA data for the average abundance of niche *g* within the 100-µm surroundings of TLSs (stable pair n = 12, relapse pair n = 10) represented as spaghetti plots (box spans interquartile range, center line indicates median, whiskers extend to 1.5× IQR) with lines connecting paired samples. Paired differences were assessed using a paired t-test, and group-by-outcome effects were assessed using an interaction test. Exact P values are reported unless P < 0.001. **(G)** Radial plot of TLS surrounding niche composition (stable pair n = 12, relapse pair n = 10).

We previously described a 26-gene immune signature (JAV-Immuno) associated with improved survival in avelumab-treated renal cell carcinoma and urothelial carcinoma cohorts (23, 24), and this signature is hypothesized to localize to TLS regions (23). In the ABACUS bulk RNA-seq dataset, baseline expression of the JAV-Immuno signature was lower in relapse samples, indicating a trend toward association with outcome at baseline in this cohort (logistic regression, p = 0.051; Supplementary Fig. 1B). Building on this observation, we next examined the spatial distribution of the JAV-Immuno signature. JAV-Immuno showed strong spatial enrichment within DIA-defined LA/TLS areas in both pre-treatment biopsies (ANOVA, p = 6.8 × 10⁻⁵) and post-treatment resections (ANOVA, p = 7.6 × 10⁻⁸) (Figure 2B, D). Publicly available TLS signatures (Cabrita-TLS) (18) similarly localized to these regions, providing orthogonal validation particularly in post samples (ANOVA, p = 1.1× 10⁻13)(Figure 2C, D). Within these TLS regions, the proportion of DIA - labeled plasma cells exhibited pharmacodynamic changes in both SD (paired t-test, p < 0.001) and relapsed (paired t-test, p = 0.0145) samples (Figure 2E).

In addition to TLS composition, the local microenvironment surrounding TLSs also showed outcome-associated differences. Specifically, a carcinoma-infiltrated stromal transition niche (niche G) was selectively reduced in SD patients post treatment (Figure 2F-G; paired t-test SD p = 0.0133; relapse p = 0.719; interaction test p = 0.0507), indicating reduced carcinoma infiltration in the stroma surrounding TLSs in SD tumors compared to relapse tumors.

Together, these data are consistent with prior reports that baseline TLS abundance is associated with anti–PD-1/PD-L1 response and demonstrate that TLS-derived transcriptional programs can be robustly detected across bulk RNA-seq and spatial transcriptomics. The availability of matched post-treatment samples in ABACUS further shows that TLS-associated transcriptional signatures are largely maintained following treatment and plasma cell composition within TLSs increase in both outcomes. In parallel, the carcinoma-infiltrated stromal transition niche adjacent to TLSs showed a borderline outcome-associated interaction. This was characterized by a reduction from pre- to post-treatment in SD tumors, indicating differences in the immediate peri-TLS microenvironment between outcomes. Together, these findings support TLSs as key immunologic structures associated with atezolizumab treatment at baseline and pharmacodynamic changes in urothelial carcinoma.

### 2.3. Atezolizumab drives broad immune and stromal remodeling

To assess broad pharmacodynamic changes associated with atezolizumab treatment, we evaluated cell classification model outputs across paired tissues. An important consideration for this analysis is that pre-treatment samples were generated from biopsies and post-treatment samples from cystectomies, which introduces inherent differences in tissue collection. Although analyses were normalized to available tissue area, we did not attempt to explicitly model harvesting related effects.

Broad pre- to post-treatment shifts were observed, including reductions in carcinoma content (logistic regression with Firth correction; OR = 0.18, p = 1.3 × 10⁻⁴) and increases in fibroblasts (OR = 8.73, p = 0.004), lymphocytes (OR = 2.70, p = 0.039), macrophages (OR = 3.24, p = 0.006), and plasma cells (OR = 2.30, p = 0.008)(Figure 3A). When stratified by clinical outcome, these changes were comparable between SD and relapse groups, with no significant outcome interaction effects in pre- versus post-treatment slopes (Supplementary Figure 1C).

**Figure 3.**
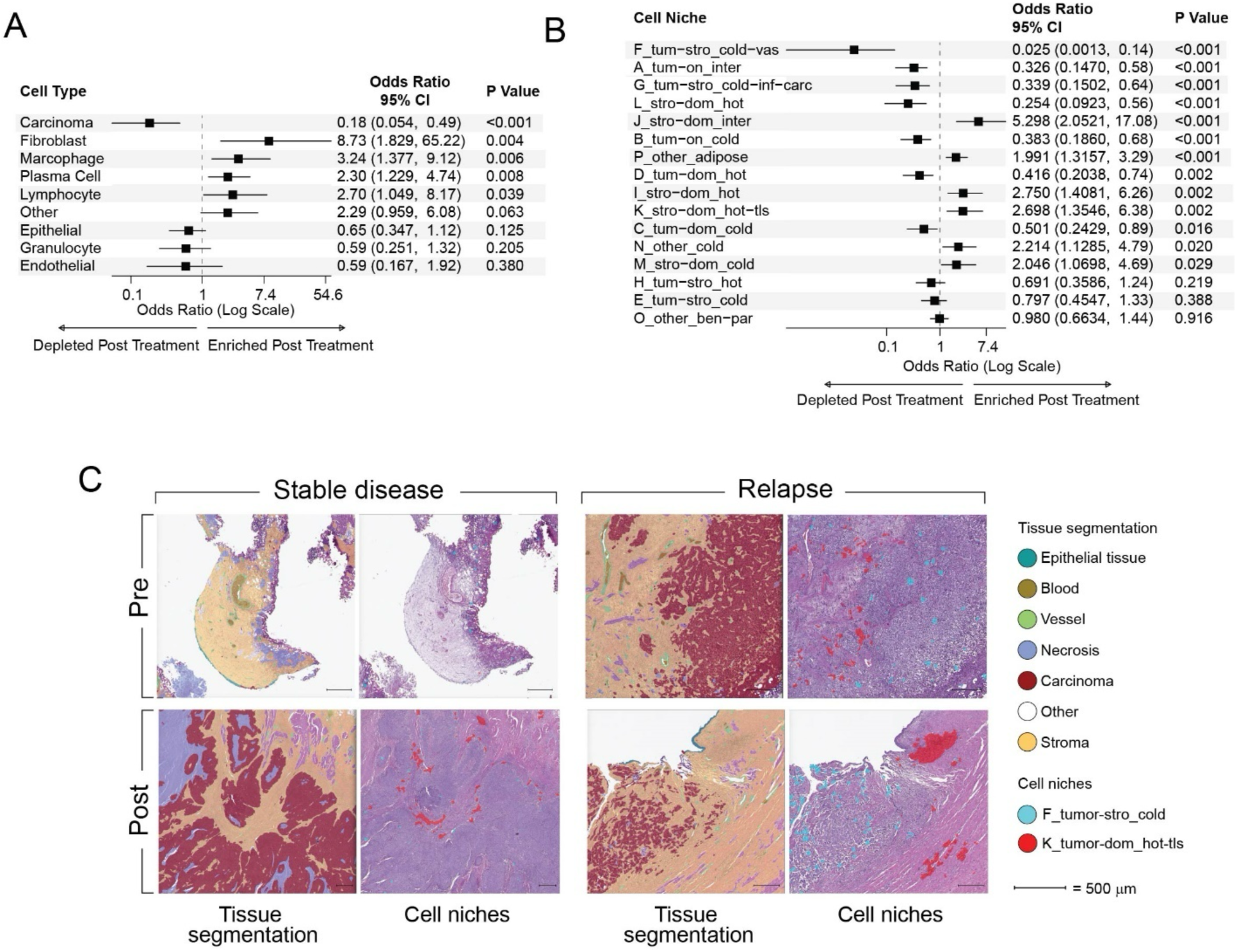
Atezolizumab drives broad immune and stromal remodeling. **(A)** Forest plot of odds ratios of cell type proportions calculated by logistic regression with Firth correction (Pre n = 26, Post n = 26). 95% CIs and p-values calculated by Profile Penalized Likelihood Ratio Test. Interaction P-values assess differences between response groups (relapse vs stable) **(B)** Forest plot of odds ratios of cell niche proportions calculated by logistic regression with Firth correction (Pre n = 26, Post n = 26). 95% CIs and p-values calculated by Profile Penalized Likelihood Ratio Test. P-values test whether enrichment differs from no change (fold change = 1). Interaction P-values assess differences between response groups(relapse vs stable) **(C)** Representative images of cell types and cell niches as overlays.

To further characterize the spatial organization of the remodeled microenvironment, we aggregated classified cells into 16 cellular niches derived from unsupervised k-means clustering followed by pathology informed merging of related clusters (25) (Supplementary Table 1). Six tumor-dominant niches decreased in prevalence, while five stromal-dominant niches increased, consistent with broad remodeling toward immune-rich and stromal-activated microenvironments. Representative examples include reduction in tumor stromal cold vasculature niche F (logistic regression with Firth correction; OR = 0.025, p = 5.34×10^-11^) and increase in stromal-dominant hot TLS niche K (OR = 2.698, p = 0.002) (Figure 3B, C). When stratified by outcome, changes in niches F and K reached significance within the SD group but not in relapse; however, no significant treatment-by-outcome interaction was detected (Supplementary Figure 1D). These results demonstrate that atezolizumab drives broad shifts in both cell type composition and higher order microenvironmental architecture, yet such global changes alone are insufficient to explain the difference between SD and relapse.

Notably, these histologic and spatial remodeling patterns closely mirror the pharmacodynamic effects reported in the original ABACUS study, which described post-treatment tumor depletion accompanied by expansion of stromal and extracellular matrix–associated programs (9). The concordance between these prior bulk transcriptomics findings and our image-based analyses supports the interpretation that the observed changes reflect canonical, treatment-associated remodeling rather than outcome-specific biology.

### 2.4. Adjacency enrichment of whole-slide images identifies differences in carcinoma-endothelial adjacency in SD vs relapse patient samples

Next, we hypothesized that carcinoma cell interactions with specific cell types may be associated with outcomes. Adjacency enrichment analysis comparing pre- and post-treatment samples revealed a significant reduction in carcinoma–endothelial interactions (mixed-effects model; OR = 0.56, p = 1.47×10^-5^), whereas carcinoma–macrophage interactions increased following therapy (OR = 1.45, p = 9.53×10^-3^) (Figure 4A).

**Figure 4.**
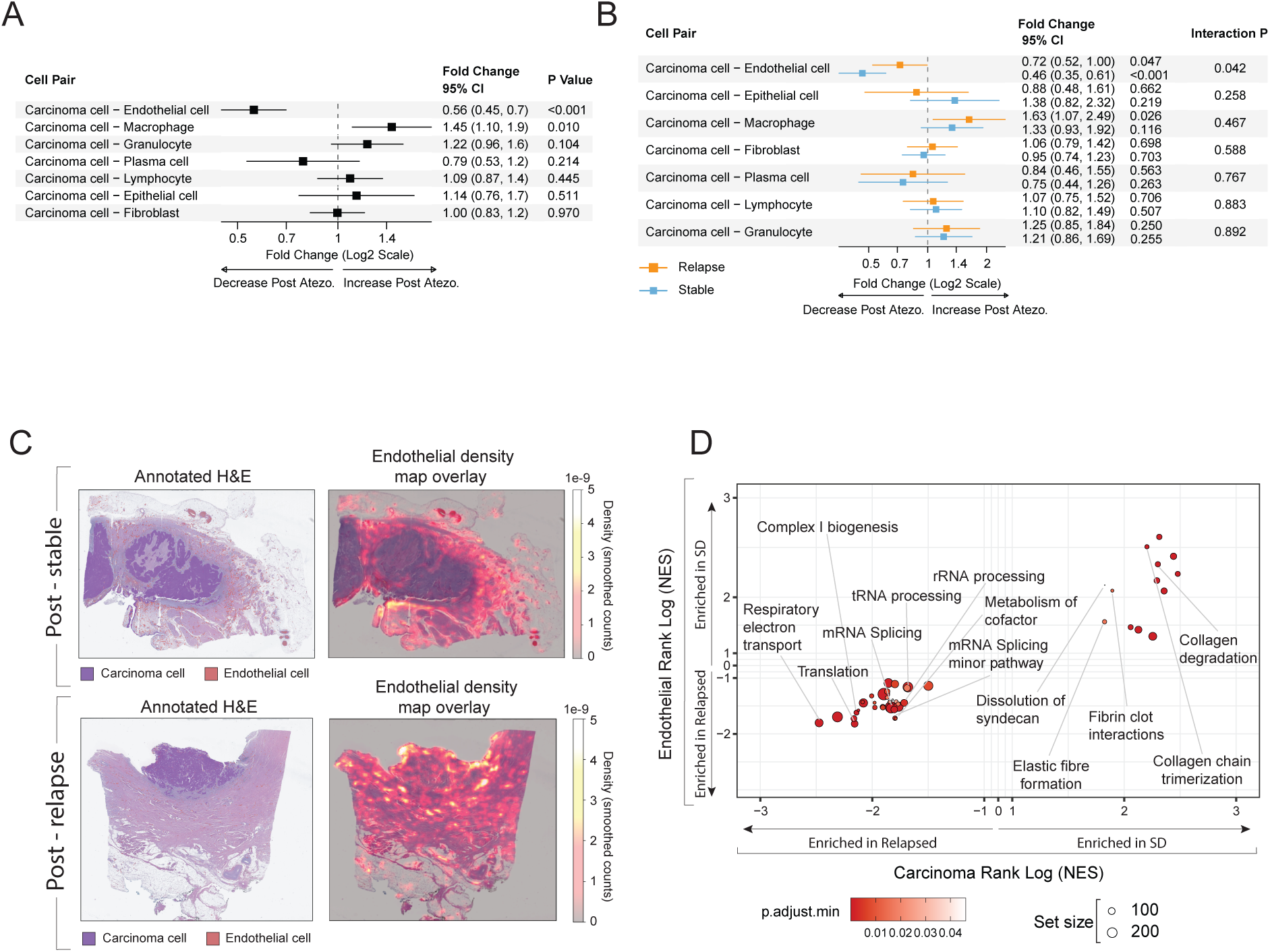
UC tumor microenvironment characterized by digital image analysis and spatial transcriptomics. **(A)** Forest plot of adjacency enrichment scores (AES) showing whole-cohort post-treatment changes in spatial adjacency between carcinoma cells and other cell types (Pre n = 26, Post n = 26). Fold changes and 95% CIs were estimated using mixed-effects models fit on log-transformed AES values. P-values test whether enrichment differs from no change (fold change = 1). **(B)** Forest plot of AES stratified by clinical outcome (stable disease n = 15; relapse n = 11). Interaction P-values assess differences in treatment-associated changes between response groups. **(C)** Representative DIA-derived spatial density maps from post-treatment samples illustrating endothelial cell density relative to carcinoma regions in stable disease and relapse. **(D)** Scatter plot of concordance between carcinoma and endothelial pathway normalized enrichment scores (NES) (stable disease pre n = 9; relapse pre n = 9; stable disease post n = 13; relapse post n = 9). Pathways were derived from rank-ordered genes and ranked using sign(ΔSD − ΔRelapse) × −log10(P), where Δ denotes post-treatment change relative to baseline.

When stratified by clinical outcome, a significant interaction was observed in the pre- to post-treatment reduction of carcinoma–endothelial proximity (interaction p = 0.042), with SD samples showing a more pronounced decrease than relapse samples (Figure 4B,C). Thus, while global treatment-associated remodeling in cell and niche proportion was similar across outcome groups, spatial decoupling of carcinoma cells from the endothelial compartment emerged as a distinguishing feature of SD.

To investigate treatment associated biological remodeling across cell compartments, we performed pathway enrichment analysis on carcinoma and endothelial-dominant spatial transcriptomic spots. Because each spatial transcriptomic spot contains multiple adjacent cell populations, spots were assigned to the most frequent cell type, generating spot level signatures that reflect the predominant, but not exclusive, cellular composition of each region. Genes were rank ordered using the interaction statistic sign (ΔSD−ΔRelapse) × −log10(interaction p) to identify pathways preferentially modulated in SD versus relapse.

Across compartments, carcinoma and endothelial regions displayed strongly correlated pathway enrichment patterns (Figure 4D). Pathways enriched in SD were dominated by extracellular matrix and adhesion processes including collagen formation, assembly of collagen fibrils, ECM proteoglycans, and integrin and nonintegrin ECM interactions. These SD-associated programs are concordant with the pronounced loss of carcinoma–endothelial adjacency observed in this group, suggesting that effective treatment is accompanied by matrix remodeling that disrupts direct tumor–vascular contact.

In contrast, relapse samples exhibited preferential enrichment of mitochondrial and translational pathways, including respiratory electron transport, aerobic respiration, complex I/III/IV assembly, fatty acid metabolism, translation, and tRNA processing. These findings indicate that tumors destined to relapse maintain active energy generating and protein synthesis programs following therapy. The persistence of these metabolic and biosynthetic pathways is consistent with the relative preservation of carcinoma-endothelial adjacency in relapse, suggesting maintenance of a supportive microenvironment that sustains tumor viability despite treatment.

### 2.5. Tumor nest border remodeling following atezolizumab

We next examined whether atezolizumab alters the cellular composition of the tumor nest (TN) border interface, reasoning that treatment-associated lymphocyte infiltration at the tumor nest border may be higher in patients with SD compared to those who relapsed. Using the H-plot approach, we generated a data-driven definition of the tumor nest border and identified a consistent zone extending ∼100 μm outward from the carcinoma boundary with 80% CI (Figure 5A).

**Figure 5.**
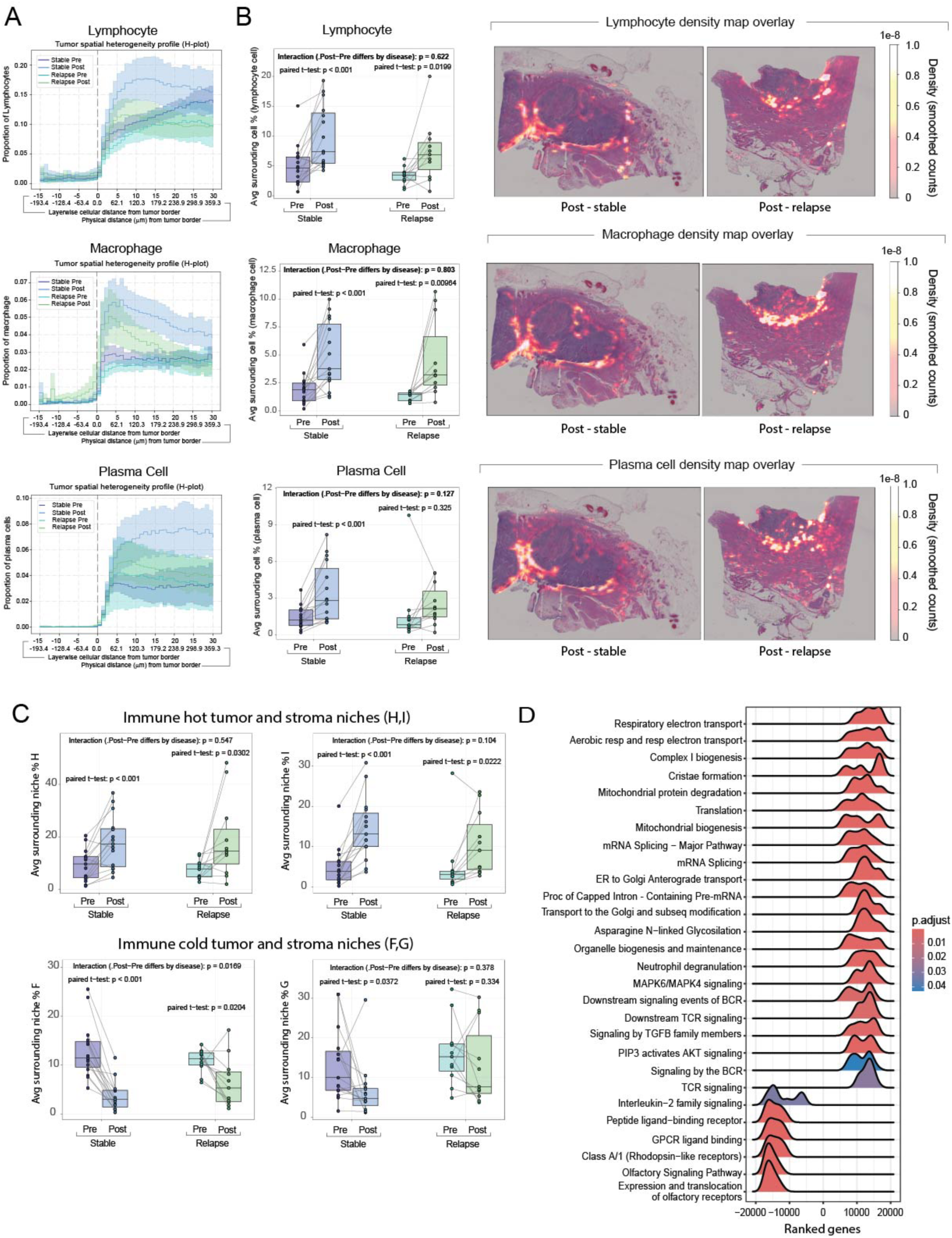
Tumor nest border remodeling following atezolizumab treatment(A) H-plots demonstrating spatial convergence of lymphocytes, macrophages, and plasma cells toward the tumor nest border and defining a conserved one hundred micrometer peritumoral zone used for all downstream analyses TN border analysis with 80% CI (stable disease pair n = 15 and relapse pair n = 11). **(B)** DIA data for the average percent lymphocytes, macrophage and plasma cell (stable disease pair n = 15, relapse pair n = 11) represented as spaghetti plots (box spans interquartile range, center line indicates median, whiskers extend to 1.5× IQR) with lines connecting paired samples. Paired differences were assessed using a paired t-test, and group-by-outcome effects were assessed using an interaction test. Exact P values are reported unless P < 0.001. Additional representative DIA-derived spatial density maps from post-treatment samples illustrating Lymphocytes, Macrophages, and Plasma cell density relative in post-treatment stable disease and relapse. **C)** DIA data for the average percent niches for niches H, I, F C G (Stable disease pair n = 15, relapse pair n = 11) represented as spaghetti plots (box spans interquartile range, center line indicates median, whiskers extend to 1.5× IQR) with lines connecting paired samples. Paired differences were assessed using a paired t-test, and group-by-outcome effects were assessed using an interaction test. Exact P values are reported unless P < 0.001. **(D)** Ridgeplot of pathways enriched in stable disease vs relapse derived from rank-ordered genes using sign(ΔSD − ΔRelapse) × −log10(P). The x-axis denotes the rank-ordered enrichment score, the y-axis shows normalized enrichment score (NES), and colors indicate pathway significance (P-value).

Across the cohort, lymphocyte (paired t-test; SD p <0.001 and relapse p = 0.0199) and macrophage (paired t-test; SD p <0.001 and relapse p = 0.00964) abundance increased within the +100 μm tumor-nest border following atezolizumab, consistent with broad treatment-associated immune remodeling at the tumor perimeter (Figure 5B). Plasma cell abundance increased significantly after treatment in SD (paired t-test; SD p < 0.001), whereas corresponding changes were not significant in relapse (paired t-test; relapse p = 0.325); however, these changes did not show consistent evidence of differential remodeling between outcomes (Figure 5B; interaction p = 0.127). Cellular-niche analysis within the 100 μm rolling-circle border region showed a similar trend, with expansion of stromal-activated and inflammatory niches (niches H and I) and a corresponding reduction in vasculature-associated cold niches (niches F and G). Reduction in Niche F reached statistical significance in the interaction between SD and Relapse in this focused TN border zone (Figure 5C).

To measure pharmacodynamic transcriptomic changes in the TN border, differential analysis was conducted between pre-treatment and post-treatment TN border spots. Results reveal increased enrichment of mitochondrial and metabolic pathways, adaptive immune receptor signaling such as downstream B cell receptor (BCR) and T cell receptor (TCR) signaling, and programs related to translation, mRNA processing, vesicular trafficking, autophagy, and TP53 regulation in post-treatment TN border regions. These changes are consistent with a metabolically activated immune state following treatment. In contrast, pre-treatment TN border regions are enriched for G-protein coupled receptor (GPCR) -mediated signaling pathways (Figure 5D). These findings demonstrate that spatially resolved profiling of the tumor nest border captures localized immune infiltration and transcriptomics activation following PD-L1 blockade that are obscured by bulk analysis.

## 3. DISCUSSION

In this study, we integrated artificial intelligence–based digital pathology with spatial transcriptomics to characterize treatment associated spatial remodeling following neoadjuvant atezolizumab in urothelial carcinoma. Using paired pretreatment biopsies and posttreatment resections from the ABACUS trial, we focused on identifying pharmacodynamic changes induced by PD-L1 blockade and assessing whether specific spatial features and their changes associate with clinical outcome.

Atezolizumab treatment induced widespread immune and stromal remodeling in both stable and relapsed disease. At the whole slide level, we observed reductions in carcinoma content alongside increases in multiple immune and stromal populations, including lymphocytes, macrophages, fibroblasts, and plasma cells. Similarly, we observed slide-level decreases in multiple tumor-stromal cold niches and increases in stromal hot niches. These global changes were consistent with prior ABACUS bulk and immunohistochemical analyses showing that immune activation and tissue remodeling are common pharmacodynamic effects of PD-L1 blockade that do not explain response heterogeneity. Similar effects have also been reported in the metastatic setting in the IMvigor210 phase II trial (10).

With spatial analyses we localized these pharmacodynamic effects to specific microenvironmental niches. LAs/TLSs were robustly detected and spatially organized relative to tumor nests. At baseline, stable disease tumors exhibited a localized enrichment of TLS-originating cells within the peritumoral region (≈200–800 µm from the tumor border) compared to relapse tumors. This intermediate peritumoral distance (200-800 µm) is consistent with the observed estimate of ∼50-200 µm for paracrine signaling by cytokines (26, 27) and it may be the minimal distance to enable immune priming near tumors.

We found that JAV-Immuno, a previously reported bulk gene expression signature of avelumab response in bladder and renal cancer, localizes specifically to LA/TLS regions (23, 24). Plasma cells, which are widely regarded as a marker of TLS maturation reflecting productive antigen presentation and adaptive immune priming (28), were increased within LA/TLSs following treatment. This maturation was further supported by coordinated changes in spatial niches surrounding TLSs, including reduced carcinoma infiltration and increased hot stroma. The observed increase in LA/TLS plasma cell proportion and nearby niches suggests that PD-L1 blockade promotes LA/TLS maturation rather than merely expanding pre-existing lymphoid aggregates.

Analyses of the TN border revealed increased lymphocyte and macrophage abundance following treatment, indicating enhanced immune access to TNs. Plasma cell abundance at the TN border increased after treatment and reached significance within the SD-group but was not detected in the relapse disease group. Transcriptomic analysis of the TN border showed a marked shift towards metabolic signatures associated with a more activated, effector state in SD patient samples. These findings suggest a potential relationship between TLS maturation and peritumoral immune activation. This is consistent with prior associations between plasma cells, anti-tumor antibody responses, CD8+ T-cell infiltration, and immune checkpoint blockade response (29–32). Altogether, these results suggest that atezolizumab induces a spatially organized immune response in multiple niches, including potential coordination between TLS maturation and peritumoral immune engagement.

While most spatial features reflected treatment associated pharmacodynamic effects that were shared across outcomes, carcinoma–endothelial adjacency emerged as a notable exception. Adjacency enrichment analysis revealed a treatment-associated reduction in tumor–vascular proximity, with a significantly greater decrease in SD compared to relapse (interaction p = 0.042). Spatial profiling revealed that atezolizumab elicits transcriptional changes in carcinoma and endothelial-dominant spots, including extracellular matrix (ECM) remodeling in SD patient samples and protein synthesis pathways in relapse patient samples. Our results are consistent with prior preclinical and clinical studies, supporting the formation of a structural barrier by abnormal vasculature and ECM organization that impairs immune cell access to tumors and their function (33–41). Reprogramming of tumor–vascular interfaces may be required for durable response to immune checkpoint blockade, and support rationale for emerging combination strategies such as PD-L1 and VEGF blocking bi-specific antibodies.

Several limitations warrant consideration. The paired design enables direct assessment of treatment associated changes but is constrained by differences between biopsy and cystectomy specimens. In addition, the modest cohort size limits power to detect moderate outcome and treatment interactions in the presence of biological heterogeneity.

As a result, many spatial features identified here should be interpreted as pharmacodynamic and hypothesis generating rather than definitive predictors of outcome.

In summary, we demonstrate how spatially aware biomarker analyses enhance understanding of mechanism of action, revealing that neoadjuvant atezolizumab induces widespread, spatially coordinated immune remodeling in urothelial carcinoma. Key novel findings of our study are that TLS maturation and increased immune infiltration with a more activated T-cell phenotype at the tumor nest border are pharmacodynamic effects of PD-L1 blockade. However, this effect was not associated with outcome, suggesting that immune infiltration and activation alone are insufficient to drive responses to atezolizumab. Reduction in carcinoma–endothelial interactions accompanied by enrichment in ECM remodeling emerged as the strongest spatial feature associated with SD. Future combination strategies inhibiting cancer-endothelial interactions may enhance response to immune checkpoint blockade.

## 4. METHODS

### 4.1. Description of Collected Samples

The ABACUS study was a single-arm phase II study to evaluate the efficacy of atezolizumab in UC in the neoadjuvant setting (9) . Between May 2016 and June 2018, 95 patients were enrolled across 21 sites. Eighty-eight patients were evaluable for the primary endpoint. Median follow-up was 13.1 months, and the median time from atezolizumab initiation to surgery was 5.6 weeks. Formalin-fixed paraffin-embedded tissue from pre-treatment biopsies and post-treatment cystectomies were utilized for HCE and Visium spatial transcriptomics.

### 4.2. Quality Control G TME analysis

Twenty-six paired hematoxylin and eosin stained whole-slide images from the ABACUS study of neoadjuvant atezolizumab in operable urothelial carcinoma were analyzed using digital pathology workflows provided by Aignostics (Berlin, Germany). Aignostics’ proprietary Atlas HCE-TME foundation model were applied sequentially and included tissue quality control (QC), tissue segmentation (TS), cell detection (CD) and cell classification (CC) models. QC excluded regions with artifacts, pen marks, and out-of-focus areas from downstream analysis. Valid tissue regions were segmented by TS into seven tissue classes (carcinoma, stroma, blood, epithelial tissue, necrosis, vessel, and other). After excluding blood and necrosis, CD identified individual cells through nucleus segmentation and CC classified those into nine cell classes (carcinoma cell, endothelial cell, epithelial cell, fibroblast, granulocyte, lymphocyte, macrophage, plasma cell, and other). The models were fine-tuned on the cohort slides and additional data at hand.

### 4.3. Cell niche clustering by neighborhood patterns

To characterize the spatial organization of classified cell types, cells were aggregated into 25 initial niches based on the composition of their individual local neighborhoods, using a k-means clustering algorithm applied to the percentages of different cell classes in 50 µm surroundings of each cell (42). The resulting niches were reviewed by a board-certified pathologist and consolidated into 16 final niches based on tissue type and immune status (supplementary table 1).

### 4.4. Tumor nest and tumor nest border characterization

TN were defined as continuous carcinoma regions and detected using TS carcinoma predictions. TN borders were defined as a 100 μm region extending outward from the TN boundary.

### 4.5. TLS detection

To detect TLSs, Aignostics developed a segmentation model identifying immature TLS (without a germinal center) and mature TLS (with a germinal center). A threshold of minimum 50 CC-determined lymphocytes per TLS was applied during post-processing to remove small lymphoid aggregates. In cases where both immature and mature regions were predicted within a single TLS, the majority class was assigned.

### 4.6. DIA Feature extraction

Based on the output of all developed models and algorithms, an integrated, comprehensive morphological feature set was extracted to characterize the spatial organization and cellular composition of the tumor microenvironment at multiple scales. At the slide level, global metrics quantified total areas and densities of regions of interest (e.g., carcinoma, stroma, epithelial tissue, vessel), cell type distributions across the nine cell classes, and the prevalence of 16 pathology-interpretable cell niches.

TLS feature extraction included categorical classification (immature vs. mature), geometric properties (area, perimeter, roundness), and cell and niche composition inside the TLS and its 100 μm surrounding. TN characterization integrated the 8-category classification system (size and embedding grade) with analogous geometric parameters, and lymphocyte infiltration metrics (maximum and average penetration depths from TN borders).

All entity-level features (TLS-wise, TN-wise) were further aggregated to slide level using summary statistics (mean, maximum, variance, sum), with separate aggregations by category.

### 4.7. Cell composition analysis

Changes in cell type and cell niche composition between pre- and post-treatment samples were assessed using logistic regression with Firth’s bias correction. Treatment timepoint (pre vs. post) was modeled as the outcome variable, with log-transformed cell composition percentages as the predictor. Firth’s penalized likelihood method was applied to address separation and small-sample bias. Odds ratios with 95% confidence intervals and P values were derived from profile penalized likelihood ratio tests.

For stratified analyses by disease status (stable vs. relapse), models were fit separately within each group. Heterogeneity of treatment effects between disease status groups was assessed by likelihood ratio tests comparing models with and without the interaction term (cell composition × disease status). Analyses were performed in R (v4.3) using the logistf package

### 4.8. H-plots

Cell classifications from DIA and TN derived from TS segmentation were utilized for tumor boundary. For each cell, the shortest Euclidean distance to the tumor boundary was computed. Cells were then assigned to discrete spatial layers based on their distance from the boundary.

Within each spatial layer, cell-type composition was quantified by aggregating cell counts across all cells assigned to that layer. For a given cell type *c* and distance layer *d*, the proportion of cells was calculated as

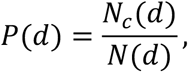

where *N*_*c*_(*d*) denotes the number of cells of type *c*in layer *d*, and *N*(*d*) denotes the total number of cells in that layer.

H-plots-were generated by plotting *P*(*d*)as a function of distance from the tumor boundary, providing a boundary-aware representation of spatial variation in cell-type composition across the tumor microenvironment.

### 4.9. Spatial adjacency enrichment analysis

Spatial relationships between cell types in the tumor microenvironment were quantified using Adjacency Enrichment Statistic (AES) (43). For each tissue sample, cell coordinates and phenotype annotations were used to construct a spatial neighborhood graph via Delaunay triangulation (44). AES values were calculated for each cell type pair by comparing the observed number of edges connecting the two cell types to the expected number under a null model of random spatial mixing. Positive values (AES > 0) indicate spatial co-localization, whereas negative values (AES < 0) indicate spatial avoidance.

Treatment-associated changes in spatial organization were assessed using linear mixed-effects models fit to log-transformed AES values for each carcinoma-associated cell pair. Visit (pre- versus post-atezolizumab) was included as a fixed effect, with patient as a random effect to account for paired samples. For response-stratified analyses, response status (SD versus relapse) and a visit × response interaction term were additionally included; interaction P values were derived from likelihood ratio tests. Fold changes from pre- to post-treatment with 95% confidence intervals were estimated from model contrasts. Analyses were performed in R (version 4.3) using lme4, lmerTest, and emmeans.

### 4.10. Visium data generation, processing G analysis

RNA was isolated from 5-micron FFPE sections using the FormaPure™ nucleic acid isolation kit (Protocol No. 000385v0005) according to manufacturer’s instructions with the addition of a DNA digestion step. RNA quality was assessed and DV200 values were determined using the Agilent TapeStation High Sensitivity RNA kit. 5-micron sections were cut on the Leica RM2235 microtome and placed on Nexterion® 3-D Hydrogel coated microscope slides, as outlined in the Visium CytAssist Spatial Gene Expression for FFPE-Tissue Preparation Guide_CG000518. FFPE samples underwent deparaffinization, decrosslinking, immunofluorescence (IF) staining and imaging as outlined in Visium CytAssist Spatial Gene Expression for FFPE_CG000520. Images were captured on the Aperio AT2 Scanner. Immunofluorescent stained slides were loaded onto the CytAssist instrument where probes were transferred onto a Visium slide. Pre-amplification was carried out and quantitation via PCR was performed on the QuantStudio7 Pro. Library preparation was performed per the manufacturers’ instructions as described in the Visium CytAssist Spatial Gene Expression for FFPE User Guide_CG000495. The final cDNA library was assessed on the Agilent TapeStation using a High Sensitivity D5000 kit prior to sequencing.

Sequencing was performed by Novogene (Sacramento, CA). Raw sequencing reads were aligned to the GRCh38 reference genome using Space Ranger v2.0.1, and feature counts were generated using the Human Transcriptome Probe Set v2.0. Post-processed Visium samples were filtered to remove samples with <1,000 median genes per spot or <0.8 confidence to the probe set. Spots with >500 detected genes, <25% mitochondrial reads, <20% hemoglobin reads, and total counts >0 were retained for downstream analysis.

For pseudo-bulk analysis, raw Visium spot level counts were aggregated across spots within each sample and tumor microenvironment category. Differential expression was assessed using DESeq2 for pre- vs post-treatment comparisons at the TN border. Carcinoma-Endothelial spots were analyzed with Data4Cure for pre- and post-stable disease versus relapse interaction analysis.

Reactome pathway analysis was performed using a signed rank score derived from differential expression results, combining log2 fold change and adjusted P-value. Gene identifiers were converted to Entrez IDs prior to analysis. Pathway enrichment was evaluated using Benjamini–Hochberg correction with a significance threshold of 0.05. Enrichment results were visualized using ridge plots and scatter plots showing pathway scores across contrasts.

## 5. STATISTICAL INFORMATION

Statistical analyses were performed using R (version 4.3). 5% false discovery rate was considered for identifying significant results, unless otherwise stated. All graphs and plots were prepared using R studio or Visual Studio Code.

## Data Availability

Data produced in the present study are available upon reasonable request to the authors

## ACKNOWLEDGEMENTS

We thank Daniel Diolaiti for editorial support and guidance on manuscript preparation. We are grateful to Paola Marcovecchio and Thomas Graham for insightful discussions on tertiary lymphoid structures and related biological context. We thank Charlotte Ackerman for coordination of ABACUS data block delivery and for facilitating access to HCE-stained images and bulk RNA-sequencing data. We also acknowledge the patients and clinical teams participating in the ABACUS study, whose contributions made this work possible.

## 6. AUTHOR CONTRIBUTION

Conceptualization: K.C., R.N., T.P., C.B.D., D.F. Methodology: K.C., R.N., M.S., C.-H.H., A.S., A.Sch., A.M.G., S.M.M., R.K., N.K.-G., S.L., C.B., M.P. Investigation: J.K., A.T., K.C., R.N., M.S., C.-H.H., A.S., A.Sch., A.M.G., S.M.M., R.K., N.K.-G., S.L., C.B., M.P., W.Y., D.C. Data analysis: K.C., R.N., M.S., C.-H.H., M.T., A.Sch., A.M.G., S.M.M. Supervision: G.L.S., G.H.Y.L., K.C., T.P., C.B.D., D.F., J.R.B., W.P. Writing original draft: R.N., G.L.S. Writing review & editing: R.N., W.P., J.R.B, G.L.S., K.C., T.P., C.B.D.

## 7. COMPETING INTEREST

T. Powles reports consulting or advisory roles with Astellas Pharma, AstraZeneca, Bristol Myers Squibb, Eisai, Exelixis, Incyte, Ipsen, Johnson & Johnson, Merck, MSD, Novartis, Pfizer, Roche, and Seagen, and travel support from Pfizer. G. H. Y. Lin, G. L. Szeto, W. Yang, D. Chawla, A. Trageser, W. Pierceall, J. Kinong, A. Sur, M. Saul, C.H. Huang, J. R. Bienkowska, D. Fernandez, K. Ching, and R. Nameki are employees of and report stock or other ownership interests in Pfizer during the conduct of this study. M. Tekman is an employee of Rancho Biosciences. M. Pezer, C. Böhm, S. Laturnus, N. Kozar-Gillan, R. Krupar, S. Munjal Mehta, A. Megina Gonzalo, and A. Schmidt are employees of Aignostics. C. B. Davis was an employee of Pfizer at the time of the study.

---

**Figure S1:**
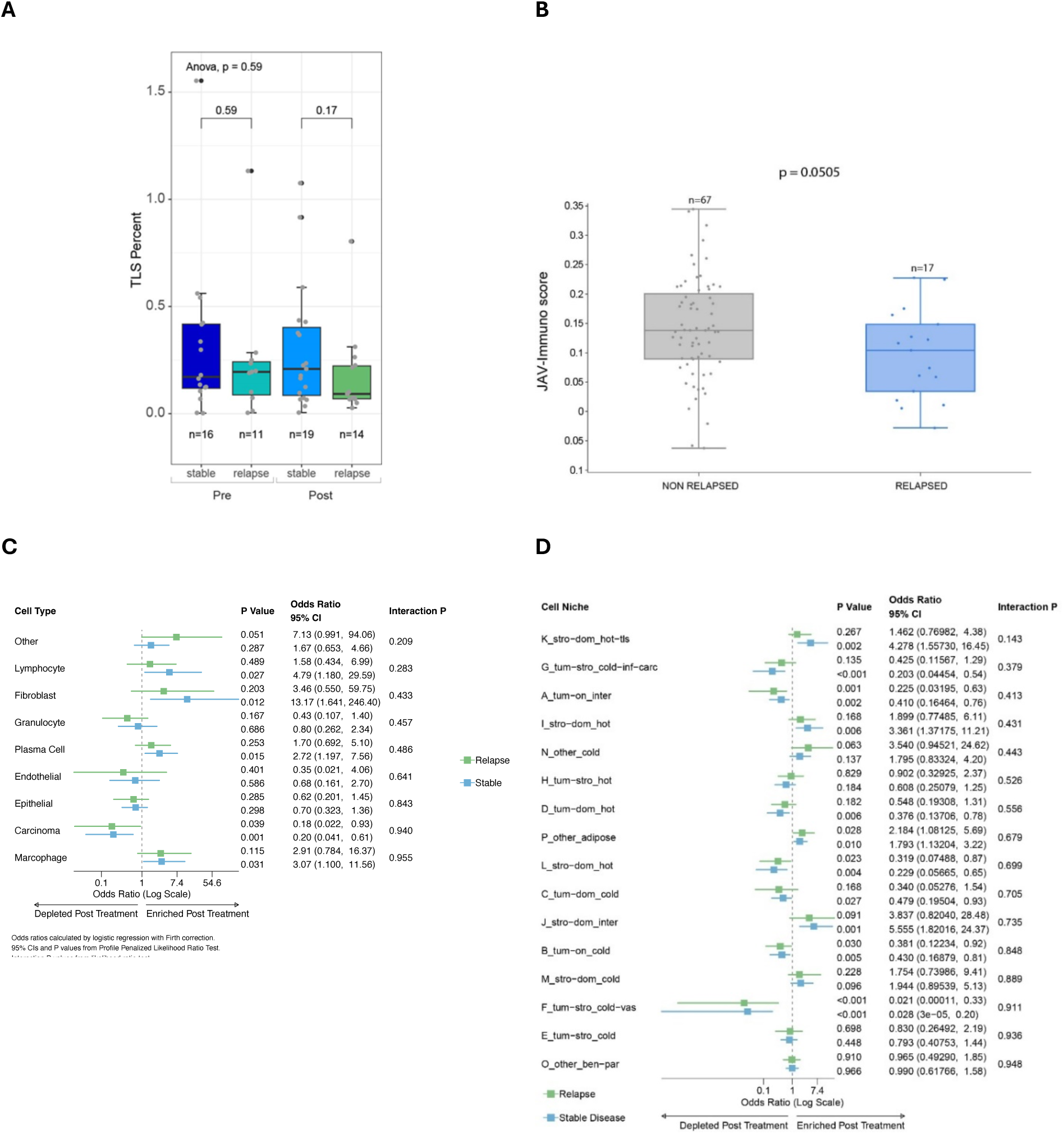
**A)** LA/TLSs (Pre stable disease n = 16, Pre relapse n = 11, Post stable disease n = 19 and Post relapse n = 14) represented as box and whisker plot (box spans interquartile range, center line indicates median, whiskers extend to 1.5× IQR). Statistical significance was assessed using unpaired t-test, with exact P values reported. **B)** JAV-Immuno score from the bulk RNA-seq from the original ABACUS biomarker study stratified by non-relapse (n = 67) and relapse (n=17) represented as box and whisker plot (box spans interquartile range, center line indicates median, whiskers extend to 1.5× IQR). Statistical significance as assessed using logistic regression, with exact P-values reported. **C)** Forest plot of pre- and post- treatment cell type and **(D)** cell niche proportions stratified by stable disease and relapse, showing similar global shifts with no significant outcome interaction.

**Table S1:**
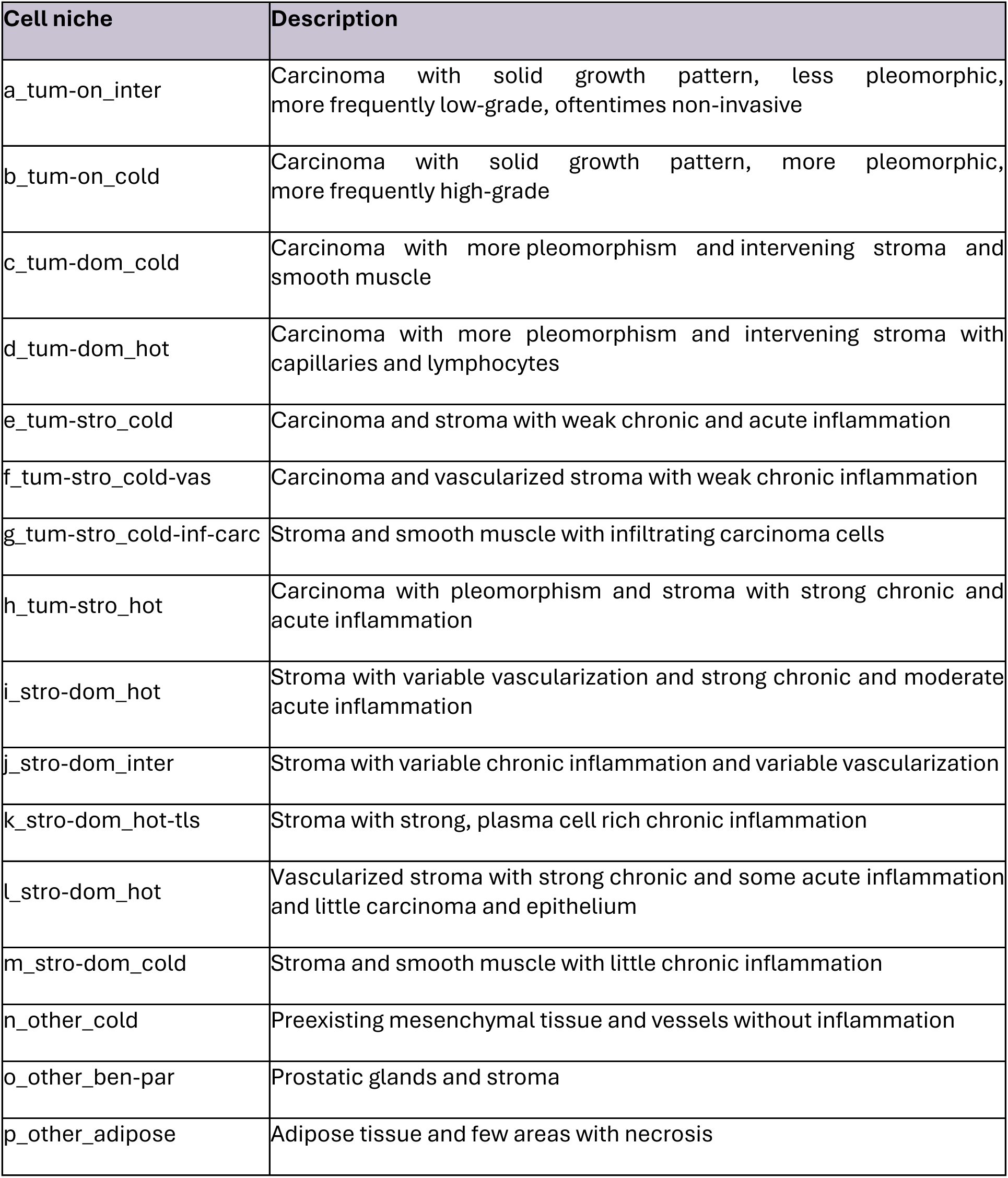
Description of 16 pathologist-consolidated cell niches.

## Notes

### Clinical Trial

NCT02662309

### Funding Statement

This work was funded by Pfizer Inc.

### Author Declarations

The present manuscript reports a secondary, retrospective analysis from the ABACUS study, a previously published multicenter, international phase II trial of neoadjuvant atezolizumab in operable urothelial carcinoma (Powles et al., Nature Medicine, 2019). All patients provided written informed consent. The relevant Institutional Review Board or Ethics Committee at each participating center approved the study, which was conducted in accordance with Good Clinical Practice and the Declaration of Helsinki. The study was sponsored and coordinated by Queen Mary University of London (Barts Cancer Institute). Ethical approval was granted. The data used had been de-identified prior to use in the study.

### Summary of Updates

Minor corrections to figures & in-line text

